# Improving the assessment of embryo developmental potential via morphokinetic forecasting of future events using language modeling

**DOI:** 10.1101/2023.10.22.23297370

**Authors:** Nir Zabari, Yoav Kan-Tor, Naama Srebnik, Amnon Buxboim

## Abstract

In IVF treatments, accurate assessment of the developmental potential of embryos to implant is essential for reaching reasonable pregnancy rates while shortening time-to-pregnancy. Hence, clinical guidelines recommend extended incubation to blastocyst transfers, which provide better evaluation of embryo developmental potential. However, cleavage stage transfer is often favored owing to various clinical considerations. To improve embryo assessment of cleavage stage embryos without extended incubation, we present a computational strategy for forecasting future morphokinetic events. Motivated by the advances in language modeling, we adapt generative pre-training to forecast future morphokinetic events based on the sequence of present events. We demonstrate < 12% forecasting error in forecasting up to three consecutive events. A new policy is proposed that combines morphokinetic forecasting and assessment of the risk of embryo developmental arrest. Using this policy, we demonstrate an improvement in the prediction of known implantation outcome of day-3 embryos from AUC 0.667 to 0.707. We expect morphokinetic forecasting to address the inherent hurdles in the selection of cleavage-stage embryos for transfer. In addition, we hope that demonstrating for the first time the utilization of language modeling on non-textual data in healthcare will stimulate future applications in reproductive medicine and other disciplines.

## INTRODUCTION

Most human embryos do not possess the capacity to implant and generate live birth. Hence, in in vitro fertilization – embryo transfer (IVF-ET) procedures, superovulation treatments are performed to eventually generate multiple embryos that will potentially serve as candidates for transfer. Maintaining reasonable pregnancy rates while shortening the time to pregnancy requires means for assessing the developmental potential of the embryos to implant and generate live birth. The utilization of time-lapse incubation systems in IVF clinics continuously generates high-quality visualization of the embryos during preimplantation development, which is exploited by various algorithms to assess embryo developmental potential^1–3^. In particular, machine learning models have been trained using the series of morphokinetic events as embryo input to predict implantation outcome. These morphokinetic features correspond to the time points that the embryo transitions between one developmental state to the next as extracted from time-lapse recordings following specific protocols. With respect to feature size, morphokinetic annotation effectively allows massive and interpretable dimensional reduction of the original video raw data (∼200 Mb per embryo). In addition, morphokinetic prediction models a-priori offer non-invasive real time developmental assessment, thus alleviating potential risks of biopsy collection and allowing fresh transfers^4^. Indeed, morphokinetic embryo assessment has been clinically tested via prospective studies and was approved for clinical applications^5^.

Recent medical guidelines recommend adopting extended incubation to single-blastocyst transfer policies worldwide. However, in many cases day-3 cleavage stage transfers are favored owing to the risk of transfer cycle cancelation due to the potential developmental arrest and lack of blastulation^6^. To increase confidence in embryo selection for transfer at cleavage stage, we hypothesized that future morphokinetic can be accurately and robustly forecasted. If so, the assessment of the potential of the embryos to implant using morphokinetic-based classifiers can be improved without extending incubation. To address morphokinetic forecasting, we realized that embryo representations via profiles of morphokinetic events is analogous to textual sentences. Specifically, the temporal associations between the morphokinetic events, which are defined by developmental constraints, are analogous to the associations between textual “tokens” and can thus satisfy the requirements for forecasting the subsequent event. This task has been actively researched and optimized, particularly during the past decade in the field of Natural Language Processing (NLP)^7–9^.

Here we employed a recently designed transformer-based auto-regressive GPT architecture, which is the current state-of-the-art NLP model. We trained this model using a multicenter dataset of 67,707 morphokinetically annotated embryos. Specifically, we used partial morphokinetic series as input (each series starts with the first developmental event) and assessed the accuracy of forecasting the next event against the existing annotated ground truth. In addition, we assess the risk of embryo developmental arrest by training a dedicated machine learning model using the morphokinetic profiles as input. Similarly, a third model was trained using embryo morphokinetic profile input to assess the potential to implant. Using these three classification models, we propose a new policy for evaluating the potential of embryos to implant which consists of the following steps: (A) Assess the potential of embryo developmental arrest. (B) For embryos with low risk of developmental arrest, forecast future morphokinetic events. (C) Evaluate the potential of the embryo to implant using the extended morphokinetic profile. Using a test set of 398 embryos, we demonstrate a significant improvement in the implantation prediction of cleavage-stage embryos three days from fertilization. In comparison with the standard policy of using the morphokinetic profiles of the recorded events (AUC 0.677), the proposed three-event forecasting policy predicted implantation outcome with AUC 0.707.

## METHODS

### Embryo database

We assembled a large multicenter database of video files of preimplantation embryo development that were morphokinetically annotated and clinically labeled as specified in a former publication^1^. In brief, the database includes a total 67,707 embryos that fertilized via intracytoplasmic sperm injection (ICSI) and were recorded on eleven time-lapse incubation systems (Embryoscope, Vitrolife) located in four medical centers. Time-lapse images were recorded with an average 18 min time interval typically for 3-to-6 days from fertilization. At each time point, seven Z-stack frames were recorded. Manual morphokinetic annotation was performed to 20,253 embryos as reported recently^1^. Five hospitals provided anonymized time-lapse video files along with corresponding metadata. Data were imported under the approval of the Helsinki ethical committee in each hospital. A qualified and experienced embryologist performed morphokinetic annotations in each IVF clinic according to established procedures^5,10^. We performed quality assurance by comparing the morphokinetic annotations of 253 randomly selected embryos with expert annotations by an embryologist in a blind manner.

To maximize the dataset size, automatic morphokinetic annotation was performed to additional 47,454 embryos. We used an automatic annotation algorithm that we recently developed with near-perfect accuracy, R-square 0.994^4^. The embryos were divided into a train-validation set (65376 embryos) and test set (2331 embryos). Importantly, all test set embryos were manually annotated using established protocols as described above.

### Prediction of embryo developmental arrest

We trained two CatBoost^11^ classification models for predicting embryo arrest at five-to-eight cells cleavage stages, and arrested morulae. Input data consists of the profile of morphokinetic events per embryo, and both models predict the potential of arrest at the most advanced input embryo state, namely the likelihood that the embryo will not reach the successive state. Since we encountered a significant imbalance between the arrested and non-arrested embryos in the test set, we randomly removed non-arrested embryos. The remaining test set included 39 five-to-eight cells cleavage stages and morula arrested embryos, and 68 non-arrested embryos. The CatBoost models were implemented using CatBoost Python library together with the default hyper-parameters. At inference time, the prediction of developmental arrest is done using one of the aforementioned classifiers, depends on the last event of the morphokinetic profile.

### Morphokinetic forecasting

Morphokinetic forecasting of embryo development was performed by training a Generative Pre-Training (GPT) model.^9^ The network architecture included 32 self-attention transformer blocks, each consisting of 32 self-attention heads. We defined the transformer token representation based on hourly rounding of embryo development from time of fertilization. For a given embryo with a given morphokinetic profile, the transformer tokens are encoded via hourly rounding of the series of morphokinetic events. The GPT model was implemented using PyTorch package with a cross-entropy loss function on predicting the next morphokinetic event.^12^ We used 65,376 train set embryos, and evaluated morphokinetic forecasting using 2,331 test-set embryos, using (normalized) mean absolute difference with varying sizes of input morphokinetic profiles. Eventually, every morphokinetic profile was enriched by 3 morphoknetic events forward using the optimized GPT.

### Embryo implantation prediction

Nowadays, implantation potential is commonly predicted using the morphokinetic profile of the embryo, via training of data-driven machine learning classifiers. As an alternative, we suggest to boost the classifier performance, by using auxiliary enrichment as a pre-processing of the training data. A forecasting model predicts one morphokinetic at most three forward, followed by an estimation of the arrest classifier. Whenever an embryo is identified as developmentally arrested, we discard it and set its probability of implantation to zero. Finally, a CatBoost model is trained with the extended morphokinetic profiles to evaluate the implantation potential.

The CatBoost model was implemented using CatBoost Python library. The hyper-parameters were selected using a grid search, implemented using sklearn^13^ Python package on the following parameters: Number of trees (taken from [30, 50, 70, 80 90, 100, 120, 140, 160]), depth of each tree (sampled from range [2,5]), and learning rate (taken from [0.01, 0.03, 0.1, 0.2]). The objective of the grid search was to select the model with the maximal AUC.

### Statistical analysis

We evaluated the statistical significance of the difference between the ROC AUC of the baseline classifier, and with our suggested policy of forecasting-arrest of 3-events forward using DeLong test^14,15^, p-value < 0.1. The 95% Confidence Interval of the ROC-AUC was (0.6-0.73) of the baseline classier and (0.63-0.77) with our suggested policy, and were calculated using Bootstrapping^15^.

## RESULTS

### Improving assessment of embryo quality by forecasting future morhokinetic events

Algorithms that predict embryo implantation outcome using a morphokinetic description of preimplantation development have demonstrated relatively high accuracy and robustness, were validated in prospective clinical trials, and gained regulatory approval. Current policy requires morphokinetic annotation of the video files that are recorded using time-lapse incubation systems according to established criteria. To eliminate inter- and intra-observed variation due to manual annotation and improve clinical workload management, algorithms that perform fully automatic annotation were recently published by us^4^ and others^16,17^. The temporal profiles of morphokinetic events are the input to implantation classification models that assess implantation potential (Fig. 1). With increasing incubation time, additional morphokinetic events can potentially add up, thus allowing the disqualification of developmentally arrested embryos and the improvement of prediction accuracy. However, various clinical considerations often require early transfer of the embryos. Here we hypothesized that assessment of the developmental potential of the embryos can be improved without extending incubation by predicting future developmental arrest and forecasting successive events (Fig. 1).

**Figure 1.**
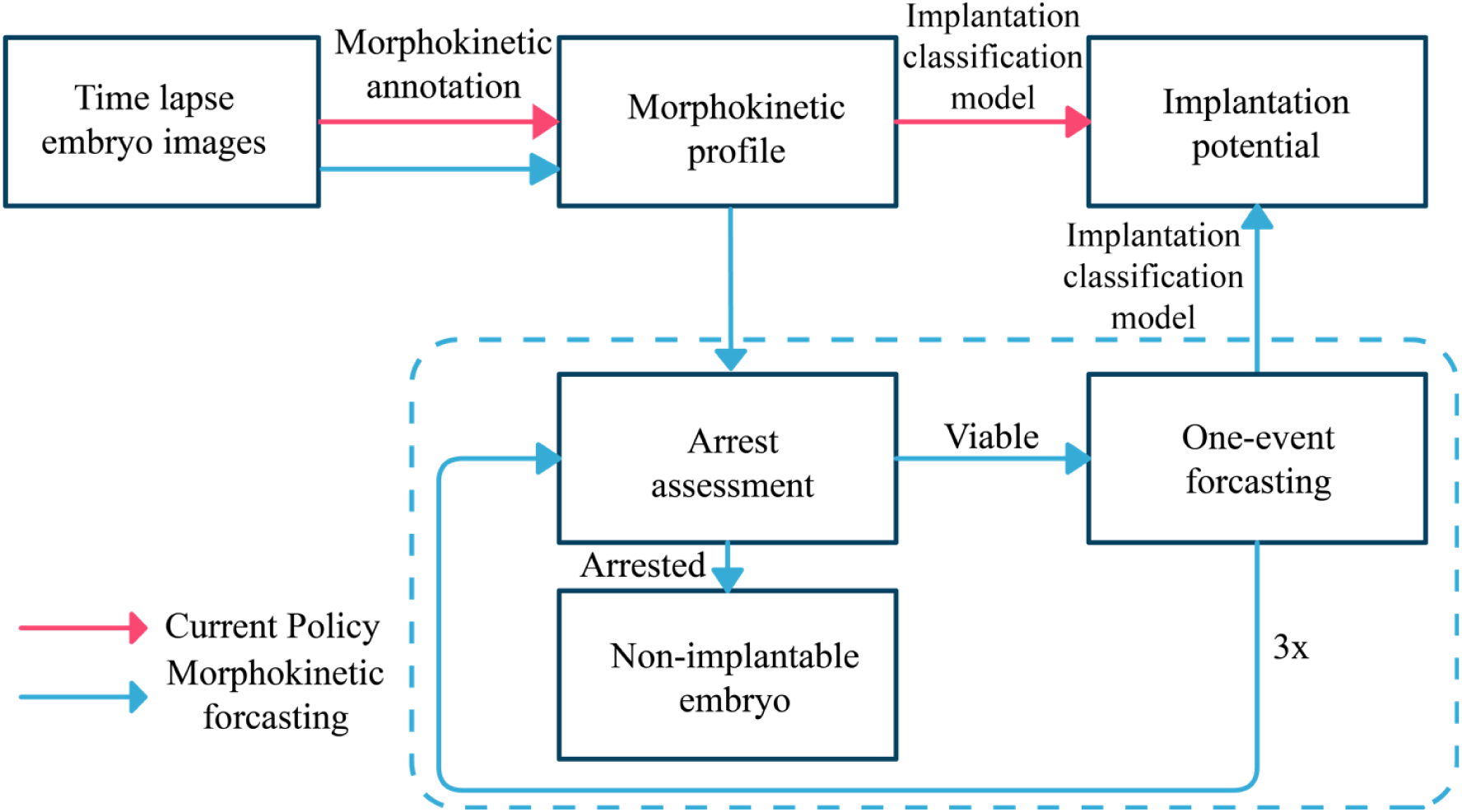
Scheme of embryo implantation prediction using extended morphokinetic profile. Current policy for implantation prediction using morphokinetic annotation of time-lapse videos is depicted in red. We propose improving implantation prediction by extending the current morphokinetic profiles via forecasting future morphokinetic events (blue). A machine-learning model is used in order to screen developmentally-arrested embryos.

### Predicting embryo developmental arrest

An arrested developmental event is defined per the most advanced embryo state reached such that advancing to the consecutive state fails despite allowing a sufficient culture time in the incubator. Hence, embryo arrest labeling requires defining the time windows for the occurrence of each morphokinetic event. To eliminate outlier contributions, we set the upper limits of these time windows to be the 97.5 percentile of the temporal distributions of the morphokinetic events. We have previously generated the most accurate statistical description of embryo preimplantation potential using 20,253 annotated embryos^4^ and used it here to calculate the temporal distributions of each event. In this manner, we were able to label arrested events as follows: An embryo is arrested at morphokinetic event *tN* if it failed to morphokinetically advance by the time of the 97.5 percentile of the t(N+1) event. Hence, labeling arrested events requires a sufficiently long recording of the embryos. Despite this limitation, we identified a total of 5,945 arrested embryos as specified in figure 2A.

**Figure 2.**
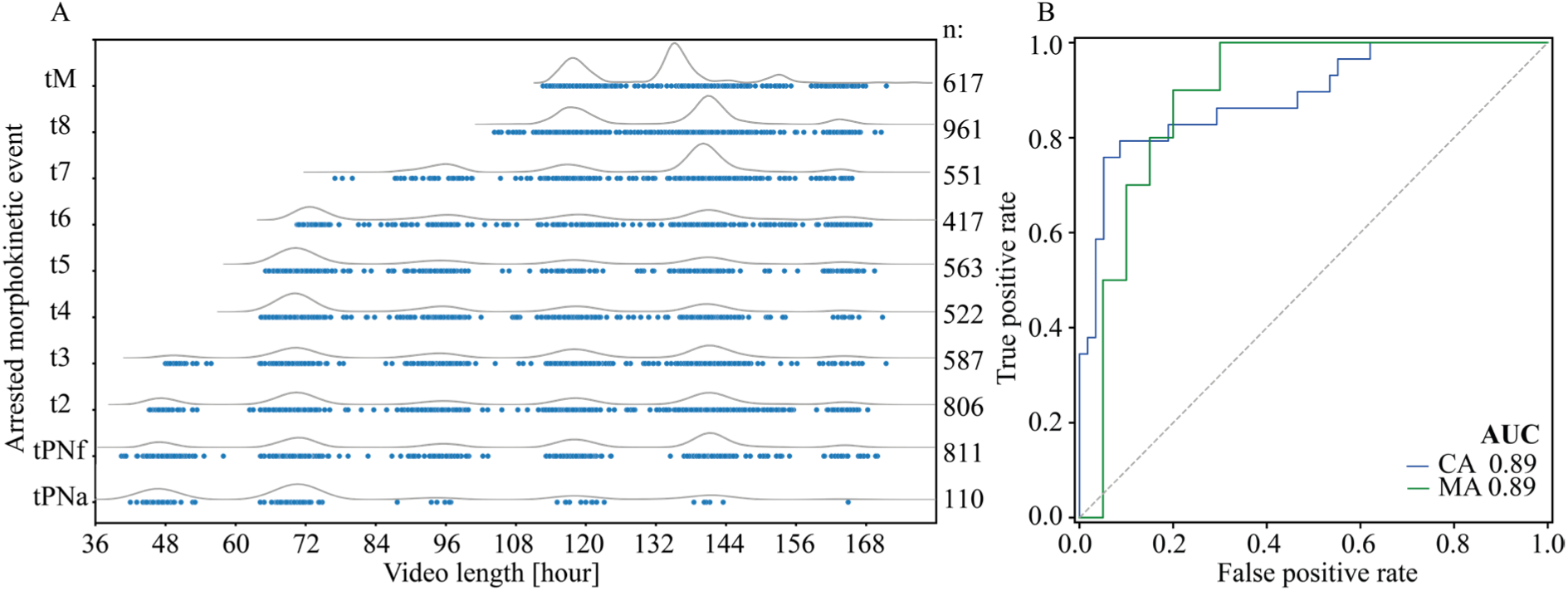
Predicting developmental arrest of embryos. **A,** The morphokinetic events at which the embryo became developmentally arrested are plotted as a function of the recorded video length. Top curves highlight the temporal densities of embryo arrest and the number of arrested embryos are shown on the right. **B,** A CatBoost model was trained using input morphokinetic profiles to predict embryo arrest at cleavage stage (CA) and at morula stage (MA). Quality of CA and MA classification models is demonstrated by the ROC curves and AUC values. ROC: Receiver operating characteristics. AUC: Area under the ROC curve.

The transfer of embryos on day-3 from fertilization or extended incubation to blastocyst transfer on day-5 are the two main transfer protocols in most IVF clinics. Embryos that reached four cells stage or earlier by day-3 are considered poor quality and are likely to be discarded. Day-3 embryos that reached six cells or more are favorable candidates for transfer. Extended incubation to blastocyst transfer is positively considered in the case that the number of high-quality day-3 embryos is sufficiently high, thus minimizing the risk of transfer cycle cancelation due to developmental arrest and lack of blastulation^6^. Consistent with current clinical practice and aiming at improving the decision on pursuing day-5 transfers, we developed classification models that predict developmental arrest at five-cells or more advanced stages. Particularly, we trained two classification models for assessing the potential of embryos to become arrested at five – to – eight cells cleavage stages and arrested morulae (Methods). We explored the prediction performance of multiple classification models and concluded that CatBoost^11^ provided highest performance. Satisfyingly, we predict embryo arrest at five – to – eight cells cleavage stages and morula arrest with AUC 0.89 (Fig. 2B).

### Embryo morphokinetic forecasting of preimplantation development

Following the assessment of arrested embryos, we set out to forecast future morphokinetic events based on the existing profiles of the morphokinetic events of the embryos, which may effectively substitute for extended incubation. To this end, we trained a forecasting Generative Pre-Training (GPT) model^9^ using a dataset of the morphokinetic profiles of 67,707 embryos (Methods). GPT models permit input morphokinetic profiles (sentences) with varying lengths as demonstrated in figure 3A. A representative embryo that was incubated for five days and reached blastulation at 86 hours from fertilization is shown. Full-range forecasting was performed using the existing profiles at t2, t4, t6 and t8 stages. We find that forecasting accuracy was improved with the length of input profiles and that time differences between forecasted and actual events were < 2 hours for cleavage stage events, < 3 hours for morula compaction, and < 4 hours for early blastulation (Fig. 3A).

**Figure 3.**
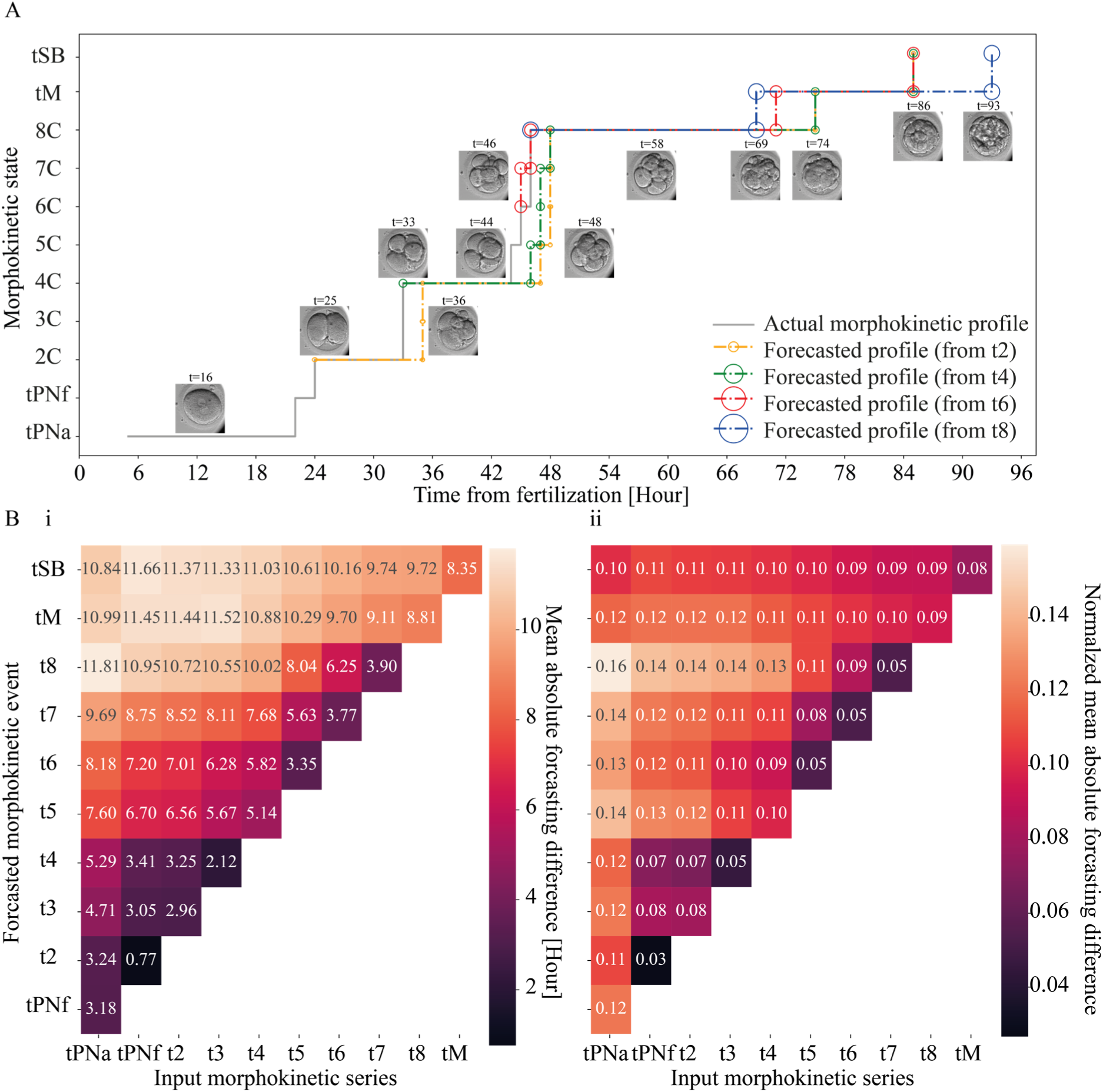
Morphokinetic forecasting of preimplantation embryo development. **A,** A staircase plot of the morphokinetic events, as defined by the discrete times of vertical developmental transitions, is demonstrated for a representative embryo (gray). Morphokinetic profiles of the same embryo were forecasted using tPNa-to-t2, tPNa-to-t4, tPNa-to-t6 and tPNa-to-t8 input events. **B,** (i) The mean absolute time differences between the forecasted and the actual morphokinetic events are shown. The rows specify the forecasted events and the columns represent the input morphokinetic profiles, from tPNa to tM. (ii) The mean absolute time differences were normalized to the time of event for each individual embryo. Forecasting errors were averaged across 1407 to 2311 test-set embryos per condition.

Next, we statistically assessed the temporal forecasting errors across the test set embryos. The mean of the absolute time differences between forecasted and actual events was evaluated for an increasing series of forecasted events using a range of forecasted input profiles from tPNa to tM (Fig. 3B-i). In parallel, we assessed the mean forecasting errors normalized to the actual time of event per each embryo (Fig. 3B-ii). We find that first cleavage-stage event forecasting error typically ranges between 2 to 4 hours accounting for 5% normalized error (with the exception of tPNf-to-t4 input profiles). tPNa-to-tPNf, tPNa-to-t3, tPNa-to-t5 and tPNa-to-t7 input profiles generated superior forecasting accuracy such that forecasting within the t2-to-t4 and t5-to-t8 blocks was more accurate than outside these blocks. Our analysis reflects the synchronized developmental blocks: Once t2 event is set, the certainty in predicting t3 is lower than predicting t2 given tPNf. Once t4 event is set, the certainty in predicting t5 is lower than predicting t4 given t3. Similarly, once t8 is set, the certainty in predicting tM is lower than predicting t6, t7 and t8 given the corresponding preceding input profiles. Within these developmental blocks, accuracy is relatively high for the forecasting of up to three-events.

### Designing an arrest-forecasting policy for improving implantation potential prediction

Current decision support tools for assessing the implantation outcome utilize various machine learning models that are based on the morphokinetic profiles of the embryos as an input^3,18–20^. Here we employed our dataset consisting of 2497 day-3 transferred embryos labeled by known implantation outcome to train a CatBoost model (Methods). Again, this classification model was chosen owing to its superior prediction performance as evaluated based on ROC-AUC metric. Our model predicts the implantation outcome of day-3 transferred embryos with AUC 0.667 (Fig. 4 black ROC curve), which is comparable with the most accurate cleavage stage classifiers.

**Figure 4.**
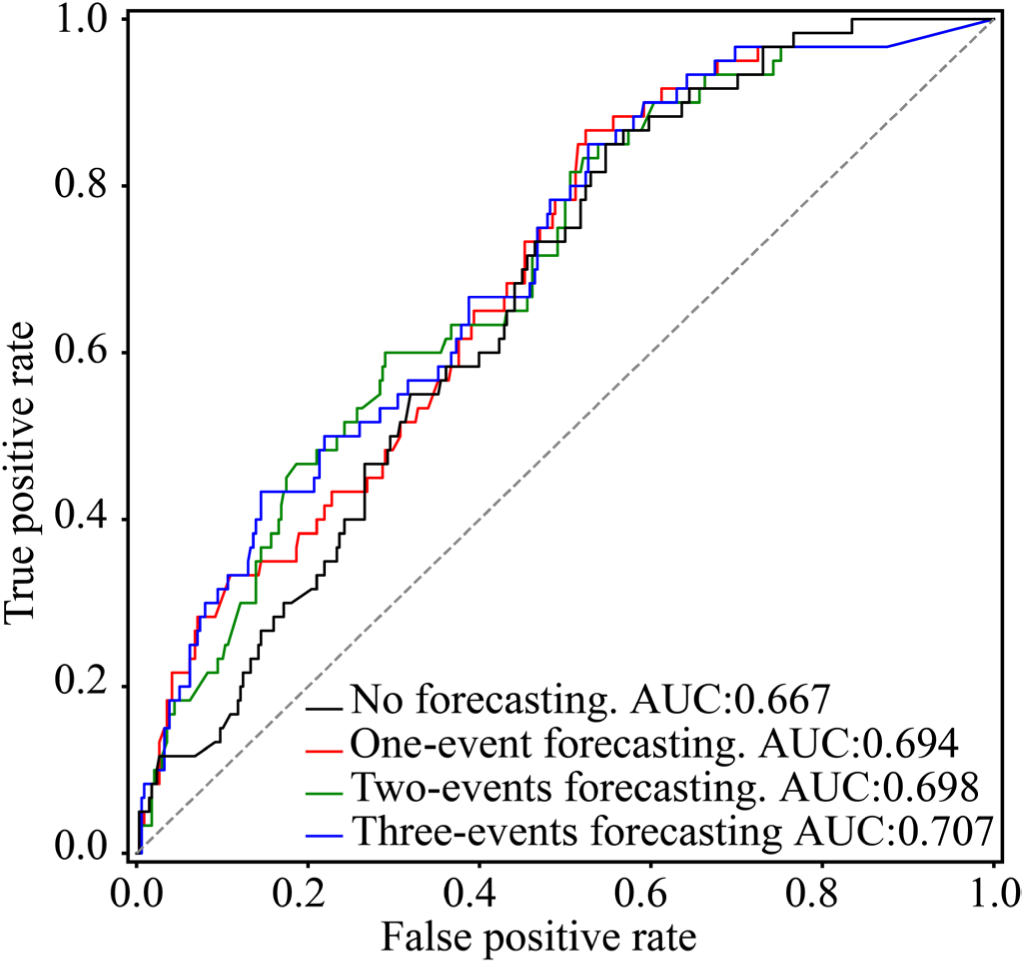
Improving embryo implantation prediction via arrest-forecasting policy. Prediction of implantation outcome was performed by training a CatBoost model using the morphokinetic profiles of the embryos at Day-3 from fertilization. The prediction of embryo implantation was improved by applying the embryo arrest – morphokinetic forecasting policy. ROC curves were evaluated for all 398 test set Day-3 transferred embryos.

Current policies for the prediction of embryo implantation outcome are based on the dimensionality-reduced representation of the available morphokinetic events as visualized by the time-lapse recordings. Here we propose to improve the prediction of implantation outcome by extending the series of morphokinetic events beyond the recorded history. The first step of the proposed policy is to evaluate the risk of developmental arrest at the current state using the classification model that is described above. This allows for deselecting the embryos that possess a high risk to become developmentally arrested. Next, we apply the GPT model for forecasting the time of the subsequent morphokinetic event. To test whether this step contributes to assessing embryo developmental potential, we applied the implantation outcome prediction model using as an input the morphokinetic series of the embryos after extending it with one forecasted event. Optionally, the developmental arrest – morphokinetic forecasting step can be repeated multiple times. With increasing the number of forecasted events, the accuracy of implantation outcome prediction is expected to improve, however morphokinetic forecasting accuracy is decreasing. To explore these contradicting contributions, we compared embryo implantation outcome prediction by the proposed policy of one, two and three event forecasting with the current no forecasting policy (Fig. 4). Current policy generated to lowest AUC score. The forecasting of one, two, and three events increased the AUC score, such that three-events forecasting demonstrated the highest prediction accuracy (AUC 0.707).

## DISCUSSION

Counter to existing guidelines recommending extended incubation to blastocyst transfer, the majority of embryo transfers to date are performed at cleavage stage owing to various clinical considerations. The selection of embryos for transfer can be guided by classification models that predict the developmental potential of the embryos to implant using different types of input data, ranging from single-frame morphology scoring^21^ to whole-video raw data^1^. In particular, morphokinetic events can serve as such input features that provide a dynamic characterization of preimplantation development and maintain low dimensionality, thus decreasing the risk of overfitting. For day-3 transfers, the morphokinetic information that can be extracted remains insufficient for accurately predicting embryo implantation outcome. As a result, day-3 fresh transfer implantation rates are significantly lower than day-5 transfers. To improve the assessment of day-3 embryos without extended incubation, we aimed at expanding the existing morphokinetic profiles by computationally forecasting future morphokinetic events. The overall goal is to contribute to accurate evaluation of embryo implantation potential, thus improving the selection of embryos for transfer to increase implantation rate.

Forecasting is a challenging problem across scientific and medical disciplines^22^. However, the advances in machine learning, the storage and processing of massive amounts of clinically labeled data, and the upscaling in computational power provide means for accomplishing forecasting tasks also in clinical settings^23–26^. Particularly, the incorporation of time-lapse incubators in IVF clinics has allowed the generation and accumulation of preimplantation videos of entire transfer cycles, including non-transferred embryos, transferred embryos, and transferred embryos with known implantation outcome labels. Standard approaches for designing machine learning classification models require the assembly of morphokinetic datasets of transferred embryos with known implantation outcome for training. However, such labeled datasets are relatively small as compared with the datasets of transferred and non-transferred embryos. In addition, there are inherent problems that are associated with manual morphokinetic annotation of a large number of embryos, including intra- and inter-observer variations and the workload resources that should be dedicated. These hurdles can be overcome using the now-available automatic morphokinetic annotation algorithms^4,16^. Akin to transfer learning approaches, we exploit the large datasets of morphokinetically-annotated datasets of non-transferred embryos and embryos that lack known implantation outcome labels for training a morphokinetic forecasting model. Forecasting is then applied to embryos with known implantation outcome.

An analogy exists between language sentences and morphokinetic sentences: (1) Morphokinetic profiles are series of discrete developmental events, just like linguistic sentences are series of discrete words. (2) In both cases, a governing mechanism exists that generates dependence between the present series and the consecutive element: biological regulatory processes of embryo development and language. (3) Despite these molecular regulatory processes and the verbal context, uncertainty exists in the identity of the consecutive element in both cases. In the past decade, significant advances in Linguistic Modeling have been achieved that provide highly accurate sentence completion^7,9^. To date, Natural Language Processing (NLP) has been implemented in healthcare only in relation with the textual domain in order to process written medical records^27,28^. Here we harnessed GPT-based modeling for performing non-textual morphokinetic forecasting. The inaccuracy that we demonstrate in forecasting up to three events is < 12% of the time of event, which is sufficient in order to improve the AUC for the prediction of embryo implantation from 0.667 to 0.707. This increase in AUC should be considered with respect to the highest reported prediction of embryo implantation outcome^19^, which did not exceed AUC 0.650.

In summary, we present a new policy for embryo selection for transfer that is based on a morphokinetic forecasting model. It combines deselecting embryos with high risk of developmental arrest and favoring the embryos with the highest potential to implant as evaluated for the extended profile with forecasted events. The clinical implementation of this policy is expected to serve as a decision support tool in IVF clinics that utilize time-lapse incubation systems. By improving assessment of embryo quality, our algorithm is expected to increase implantation rates while shortening time-to-pregnancy. Finally, to the best of our knowledge, we demonstrate for the first time the utilization of Language Modeling of non-textual data in healthcare, which we hope will stimulate future work in reproductive medicine as well as in other medical fields.

## DECLARATIONS

## Competing interests

N.S., M.G., U.S., and A.B. declare no financial or non-financial competing or other conflict of interest.

Y.K.T. declares no financial or non-financial competing or other conflict of interest. Since September 2021, Y.K.T. discloses being employed by IBM-Research.

## Ethical approval

This research was approved by the Investigation Review Boards of the data-providing medical centers: Hadassah Hebrew University Medical center IRB number HMO 558-14; Kaplan Medical Center IRB 0040-16-KMC; Soroka Medical Center IRB 0328-17-SOR; Rabin Medical Center IRB 0767-15-RMC.

## Code and data availability

The copyrights of the code are owned by Yissum–the technology transfer company of The Hebrew University of Jerusalem. Requests can be sent to A.B. The clinical data are owned by Hadassah Medical Center and by Clalit Health Services. Restrictions apply to the availability of these data, which were used anonymously under ethical agreements with each clinic separately for this study, and so are not made publicly available.

## Funding

A.B. greatly appreciates support from the European Research Council – Proof of Concept Grant (PoC 966830) and the European Research Council Grant (ERC-StG 678977). The funders did not play any role in study design, data collection and analysis, decision to publish, or preparation of the manuscript.

## Data Availability

The copyrights of the code are owned by Yissum which is the technology transfer company of The Hebrew University of Jerusalem. Requests can be sent to A.B. The clinical data are owned by Hadassah Medical Center and by Clalit Health Services. Restrictions apply to the availability of these data, which were used anonymously under ethical agreements with each clinic separately for this study, and so are not made publicly available.

## Notes

### Competing Interest Statement

Competing interests
N.Z., N.S. and A.B. declare no financial or non-financial competing or other conflict of interest.
Y.K.T. declares no financial or non-financial competing or other conflict of interest. Since September 2021, Y.K.T. discloses being employed by IBM-Research.

